# Upadacitinib and Cardiovascular Adverse Events in Rheumatoid Arthritis: A Systematic Review and Meta-Analysis

**DOI:** 10.1101/2023.10.24.23297509

**Authors:** Fatemeh Omidi, Parisa Delkash, Tala Sarmastzadeh, Mohammad Javad Nasiri

**Affiliations:** Department of Cardiology, Imam Hossein Hospital, Shahid Beheshti University of Medical Sciences, Tehran, Iran; Department of Rheumatology, Imam Hossein Hospital, Shahid Beheshti University of Medical Sciences, Tehran, Iran; School of Medicine, Shahid Beheshti University of Medical Sciences, Tehran, Iran

**Author notes:** Correspondence Parisa Delkash, MD, Mohammad Javad Nasiri, PhD, MPH.

**Keywords:** Upadacitinib, Janus kinase inhibitor, cardiovascular adverse events, rheumatoid arthritis, meta-analysis

## Abstract

**Background:** The safety of Upadacitinib, a Janus kinase inhibitor, in the context of rheumatoid arthritis management has raised concerns regarding potential cardiovascular adverse events, but the evidence remains inconclusive.

**Methods:** Our study involved a systematic search for articles conducted up to October 1, 2023, encompassing databases such as PubMed/Medline, Embase, and Cochrane CENTRAL. We employed meta-analysis to calculate pooled odds ratios (OR) and their 95% confidence intervals (CI). We assessed potential publication bias through the application of Begg’s and Egger’s tests.

**Results:** Six studies involving 4202 patients were included. The analysis of the 15 mg dosage revealed a pooled OR of 1.20 (95% CI: 0.3-4.3), indicating a small increase in cardiovascular adverse event likelihood without statistical significance. The 30 mg dosage analysis yielded a combined OR of 2.37 (95% CI: 0.6-9.1), suggesting a potential risk increase but lacking statistical significance. Begg’s and Egger’s tests indicated no publication bias.

**Conclusion:** While there is a suggestion of elevated cardiovascular risk, especially with the 30 mg dosage, the absence of statistical significance and wide confidence intervals underscore the need for cautious interpretation. Individualized treatment decisions, vigilant monitoring, and further research are essential to optimize patient care and deepen our understanding of Upadacitinib’s safety profile.

## Introduction

Rheumatoid arthritis is a chronic autoimmune disease characterized by joint inflammation, and pain [1]. This condition, if left uncontrolled, can lead to joint damage, disability, reduced quality of life, and potentially, cardiovascular and other associated complications [1-3]. The global incidence of rheumatoid arthritis remains relatively stable at approximately 0.5% to 1.0%, with a higher prevalence among women and the elderly [4, 5]. As the treatment landscape for rheumatoid arthritis evolves, the introduction of Janus kinase inhibitors, such as Upadacitinib, has promised enhanced symptom management and improved quality of life for patients [6-8]. Nevertheless, the emergence of cardiovascular adverse events as a potential side effect of JAK inhibitors has raised crucial questions about their overall safety profile [9-13]. Indeed, while some studies have examined the safety profile of Upadacitinib, there is a recognized need for a comprehensive and up-to-date investigation [14-16]. In response to these concerns, we present a robust systematic review and meta-analysis that rigorously examines the relationship between Upadacitinib use and cardiovascular adverse events in patients diagnosed with RA.

## Methods

The present study was performed and reported following the PRISMA guideline (Pending ID: 475681) [17].

### Search strategy

Medical databases including PubMed/Medline, EMBASE, and Cochrane CENTRAL were searched for relevant studies published up to October 20, 2023. Only RCT written in English were selected. We used the following combinations of the MeSH terms and keywords: Janus Kinase Inhibitors, Upadacitinib, and rheumatoid arthritis. Backward and forward citations were searched in selected studies to identify further relevant publications.

### Study Selection

The data collected from various sources were consolidated, and any redundant entries were eliminated through the utilization of EndNote X8. Each record was independently screened by two reviewers (M.N or T.S) for eligibility criteria and excluded unrelated studies based on title/abstract, followed by full text. In case of a discrepancy between the two reviewers, the lead investigator assessed the record. Eligible studies met the following criteria:

The inclusion criteria were:

*Study design:* randomized controlled trials (RCTs)

*Patients:* With a clinical diagnosis or symptoms based on the American College of Rheumatology (ACR) Guideline.

*Interventions:* Upadacitinib

*Comparisons:* Placebo

*Outcomes:* Cardiovascular events

Conference abstracts, editorials, reviews, expert opinions, duplicate studies, study protocols, case reports, and case series were excluded.

### Data extraction

Two reviewers (M.N or MR) designed a data extraction form and extracted data from all included studies. The data of each record were extracted by two reviews and differences were resolved with consensus. The following data were extracted: first author names; year of publication; study design; mean age; follow up; number of participants; interventions; control, and outcomes.

### Quality assessment

The quality of each study was assessed by two reviewers (M.N, T.S) and a third reviewer was involved to resolve any inconsistencies. Items such as study population, sampling, methods of identification and measure of the condition, and statistical analysis were evaluated by using the Cochrane bias assessment tool.

### Data analysis

We conducted statistical analyses using Comprehensive Meta-Analysis software, version 3.0, developed by Biostat Inc., located in Englewood, NJ, USA. Pooled odds ratios (OR) were calculated, along with their corresponding 95% confidence intervals (CIs), employing random-effects models in conjunction with the Mantel-Haenszel statistical approach. To gauge between-study heterogeneity, we employed Cochran’s Q test and the I2 statistic. We assessed potential publication bias by employing Begg’s and Egger’s tests, with a significance threshold of P < 0.05 indicating the presence of statistically significant publication bias.

## Result

From our initial database searches, we identified a total of 2140 citations. Following a screening process that involved assessing titles and abstracts, we retrieved full-paper copies for 49 citations that appeared to be potentially suitable for inclusion in our review. Subsequently, we excluded 43 full-text studies, as delineated in Figure 1. Ultimately, we incorporated the remaining 6 RCTs comprising 4202 participants into our analysis, as they met the established minimum criteria for inclusion [18-23].

**Figure 1.**
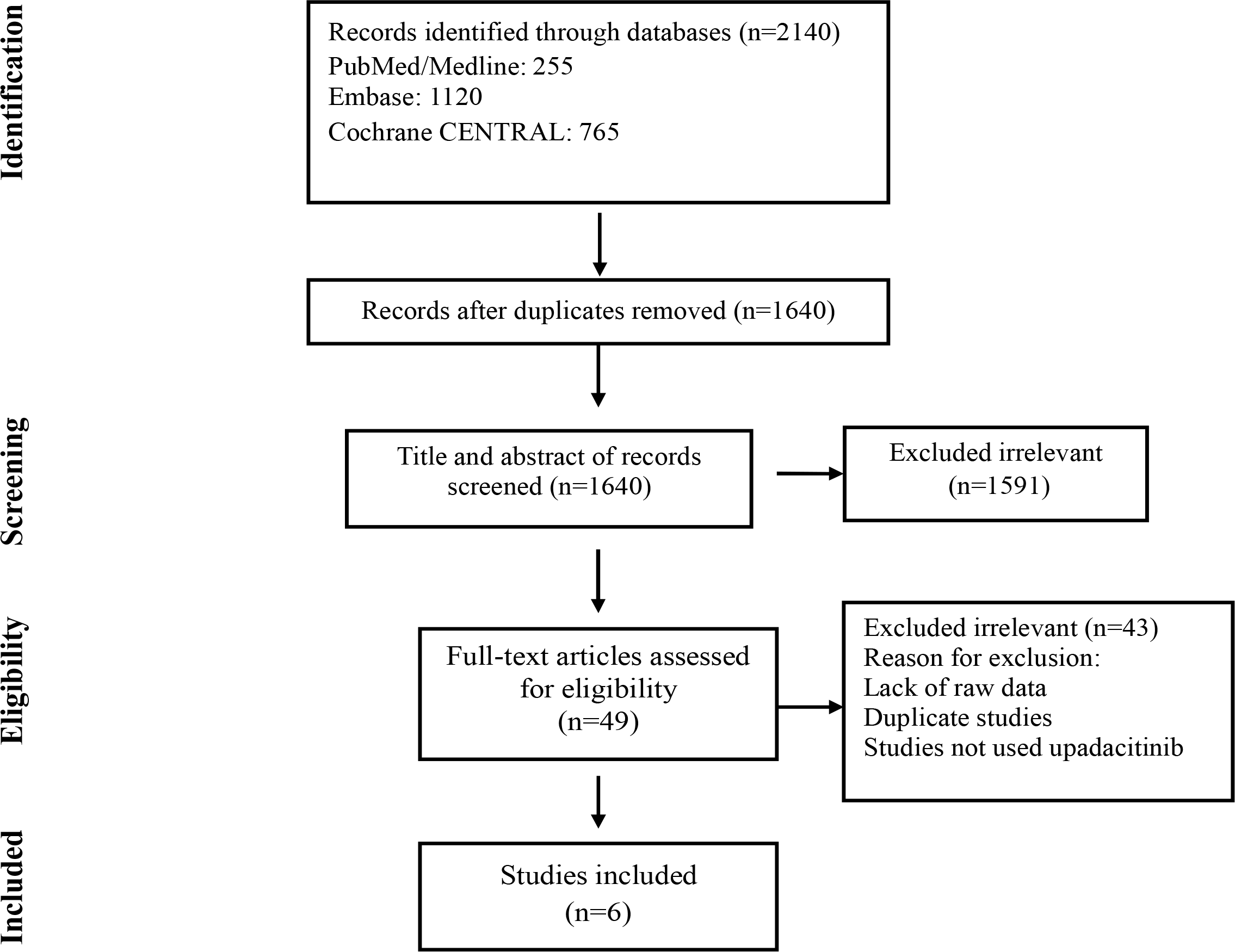
Flow chart of study selection for inclusion in the systematic review and meta-analysis

### Study characteristics

Table 1 provides an overview of study characteristics, including details about the study setting, design, participant count, average age, applied criteria, and follow-up duration. Six randomized controlled trials investigated the effectiveness of upadacitinib as a treatment for rheumatoid arthritis. These trials generally had a duration of 12 to 24 weeks, with sample sizes ranging from around 200 to 650 patients. The mean age of patients across the studies was typically in the range of 50 to 55 years. In each study, patients with rheumatoid arthritis were divided into groups receiving upadacitinib and placebo as a control. The criteria for assessing rheumatoid arthritis in these studies were based on the American College of Rheumatology (ACR) guidelines.

**Table1.** Studies characteristics.

| **First Author, Year** | **Study design/Duration** | **Sample size Upadacitinib/ control** | **Type of patient, Mean age** | **Intervention** | **Control** | **Criteria for rheumatoid arthritis** |
| --- | --- | --- | --- | --- | --- | --- |
| Burmester-2018 [18] | RCT/12 weeks | 440/221 | Patients with rheumatoid arthritis, 55 | Upadacitinib 15 and 30mg qd | Placebo | ACR |
| Genovese-2018 [19] | RCT/12 weeks | 329/169 | Patients with rheumatoid arthritis, 50 | Upadacitinib 15 and 30mg qd | Placebo | ACR |
| Fleischmann-2019 [20] | RCT/24 weeks | 650/652 | Patients with rheumatoid arthritis, 54 | Upadacitinib 15mg qd | Placebo | ACR |
| Smolen-2019 [21] | RCT/12 weeks | 432/216 | Patients with rheumatoid arthritis, 54 | Upadacitinib 15 and 30mg qd | Placebo | ACR |
| van Vollenhoven-2020 [22] | RCT/24 weeks | 631/314 | Patients with rheumatoid arthritis, 52 | Upadacitinib 15 and 30mg qd | Placebo | ACR |
| Kameda-2020 [23] | RCT/12 weeks | 99/49 | Patients with rheumatoid arthritis, 55 | Upadacitinib 15 and 30mg qd | Placebo | ACR |

### Risk of bias assessment

As outlined in Table 2, our risk of bias assessment indicated that the included studies generally demonstrated a low risk of bias across the critical domains. These domains encompass critical criteria, such as randomization procedures, blinding of both participants and assessors, the completeness of outcome data, selective reporting, and potential sources of bias.

**Table 2.** Quality Assessment.

| Author | Random<br>Sequence<br>Generation | Allocation<br>Concealment | Blinding Of<br>Participants<br>And<br>Personnel | Blinding Of<br>Outcome<br>Assessment | Incomplete<br>Outcome<br>Data | Selective<br>Reporting |
| --- | --- | --- | --- | --- | --- | --- |
| Burmester | Low Risk | Low Risk | Low Risk | Low Risk | Low Risk | Low Risk |
| Genovese | Low Risk | Low Risk | Low Risk | Low Risk | Low Risk | Low Risk |
| Fleischmann | Low Risk | Low Risk | Low Risk | Low Risk | Low Risk | Low Risk |
| Smolen | Low Risk | Low Risk | Low Risk | Low Risk | Low Risk | Low Risk |
| van Vollenhoven | Low Risk | Low Risk | Low Risk | Low Risk | Low Risk | Low Risk |
| Kameda | Low Risk | Low Risk | Low Risk | Low Risk | Low Risk | Low Risk |

### Cardiovascular Adverse Events with upadacitinib 15 mg

The occurrence of cardiovascular adverse events resulting from the use of 15 milligrams of upadacitinib was investigated in six studies. The pooled OR was found to be 1.20 (CI 95%: 0.3-4.3, I2: 0.00). This odds ratio suggests a relatively small increase in the likelihood of experiencing cardiovascular adverse events in the upadacitinib group compared to the control group. However, the odds ratio was not statistically significant and do not provide strong evidence to conclude that the use of upadacitinib significantly increases the risk of cardiovascular adverse events (Figure 2). Based on the results of the Begg’s and Egger’s tests, there was no indication of publication bias (p>0.05).

**Figure 2.**
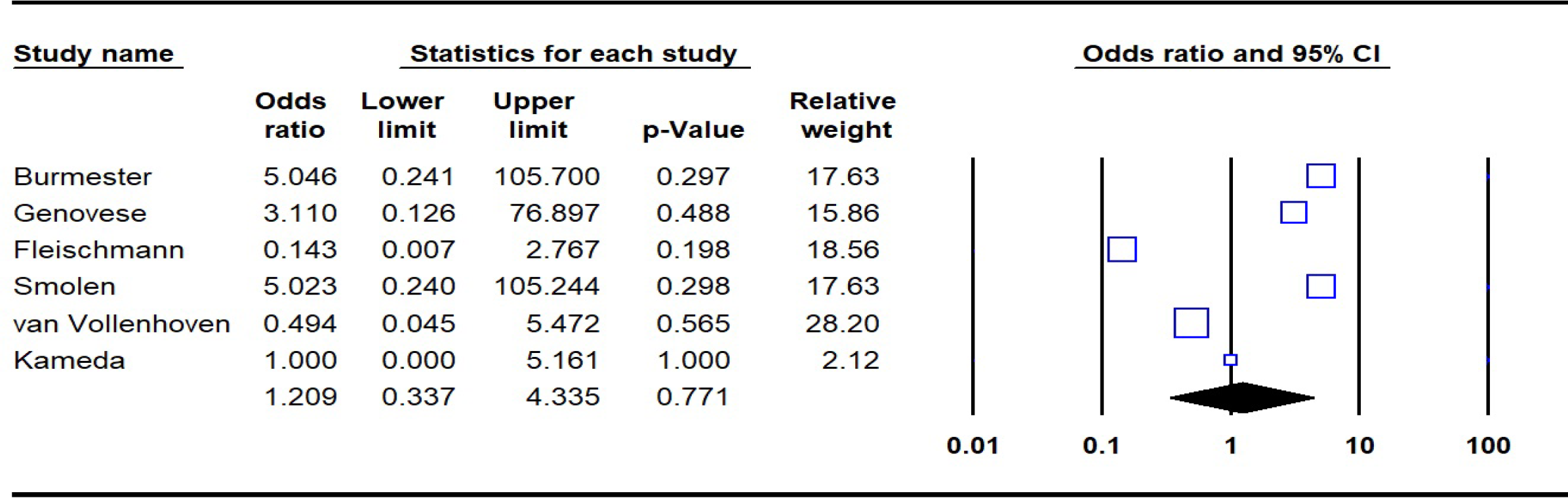
Pooled OR of Upadacitinib 15 mg versus placebo

### Cardiovascular Adverse Events with upadacitinib 30 mg

The investigation into cardiovascular adverse events associated with the use of 30 milligrams of upadacitinib encompassed five studies. The analysis yielded a combined OR of 2.37, with a 95% CI ranging from 0.6 to 9.1. The low I2 statistic value (I2: 0.00) indicates minimal variability among the studies. While the point estimate suggests a potential increase in the risk of cardiovascular adverse events in the upadacitinib group, the broad confidence interval prevents this estimate from being statistically significant (Figure 3). Furthermore, the results from both Begg’s and Egger’s tests indicated an absence of publication bias (p>0.05).

**Figure 3.**
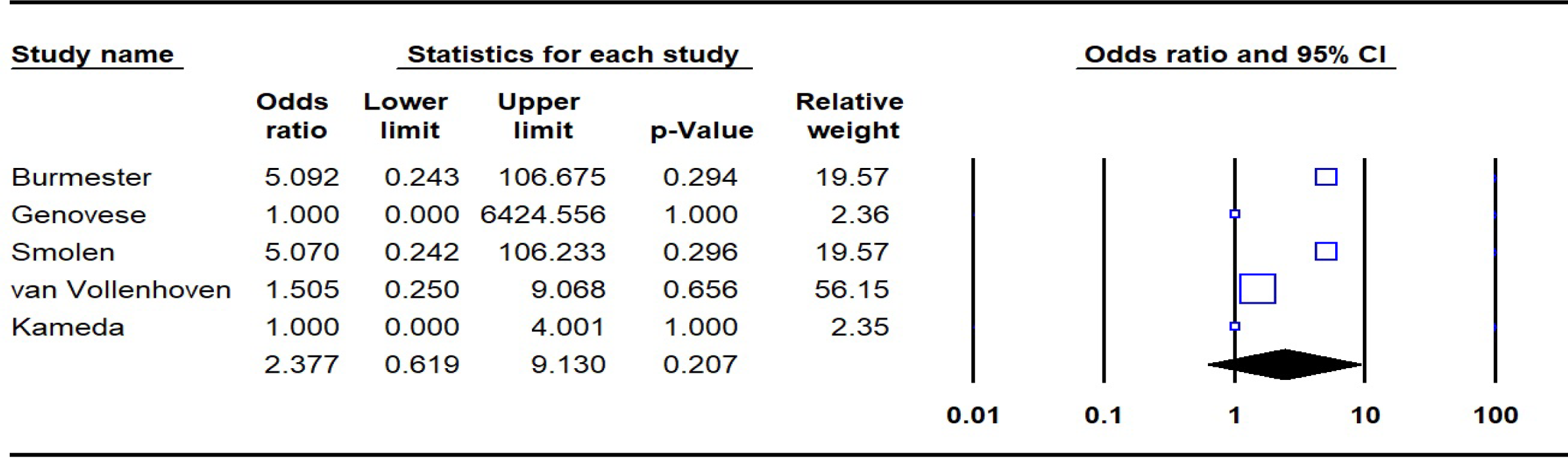
Pooled OR of Upadacitinib 30 mg versus placebo

## Discussion

Assessing cardiovascular adverse events linked to two distinct dosages of Upadacitinib, offers valuable insights regarding the safety profile of this medication within the context of rheumatoid arthritis management. Also, the observed estimate suggests a relatively modest increase in the likelihood of experiencing cardiovascular adverse events in the Upadacitinib groups, however, our findings do not provide robust evidence to conclude that the use of Upadacitinib significantly elevates the risk of cardiovascular adverse events.

### Clinical Implications

The results derived from our systematic review and meta-analysis offer important insights for clinicians, researchers, and healthcare policymakers in the context of managing rheumatoid arthritis with Upadacitinib.

#### Individualized Treatment Decisions

Clinicians should consider these findings when making individualized treatment decisions for rheumatoid arthritis patients [24-26]. While Upadacitinib offers therapeutic benefits in managing rheumatoid arthritis symptoms, the potential for cardiovascular adverse events should be weighed against these benefits. Patient-specific factors, including existing cardiovascular risk factors, should be carefully assessed when prescribing Upadacitinib.

#### Regular Cardiovascular Monitoring

Patients on Upadacitinib therapy, especially those receiving the 30 mg dosage, should undergo regular cardiovascular monitoring. This includes assessing cardiovascular risk factors, conducting electrocardiogram, and tracking blood pressure and lipid profiles [27-30]. Early detection and intervention can help mitigate potential cardiovascular risks.

#### Alternative Treatment Options

In cases where the potential cardiovascular risks associated with Upadacitinib are a concern, clinicians may consider alternative treatment options for rheumatoid arthritis management. The choice of therapy should be tailored to the individual patient’s needs and risk profile.

### Strength and limitation

#### Strengths

##### Comprehensive Analysis

Our study includes a comprehensive analysis of a substantial number of studies, encompassing a significant sample size. This provides a robust overview of the safety profile of Upadacitinib in patients with rheumatoid arthritis.

##### Assessment of Multiple Dosages

We assess the safety of two different dosages of Upadacitinib, 15 mg, and 30 mg. This approach enables a nuanced understanding of potential dose-dependent effects on cardiovascular adverse events.

#### Limitations

##### Wide Confidence Intervals

The wide confidence intervals observed in our analysis indicate a degree of uncertainty around the point estimates. This uncertainty may be due to variations in study populations, designs, and other factors.

##### Lack of Long-Term Data

Many of the studies included in our analysis are relatively short in duration. Long-term data on the safety of Upadacitinib, especially with extended use, are limited.

##### Potential Confounders

Our analysis is based on available data from the included studies. We cannot account for potential confounding variables that may influence cardiovascular adverse events, such as individual patient characteristics and concomitant medications.

In conclusion, our systematic review and meta-analysis offer insights into the safety of Upadacitinib in rheumatoid arthritis management. While our findings suggest a potential increase in cardiovascular adverse events, particularly with the 30 mg dosage, the lack of statistical significance and wide confidence intervals underscore the need for cautious interpretation. These results align with concerns about Janus kinase inhibitors’ safety, emphasizing the importance of individualized treatment decisions, vigilant monitoring, and shared decision-making with patients. Further research, robust clinical trials, and real-world evidence are necessary to deepen our understanding of Upadacitinib’s safety profile.

## Data Availability

All data produced in the present work are contained in the manuscript

## Author contribution

All authors contributed to the final manuscript.

